# Clinical Effects of Physiologic Lesion Testing in Influencing Treatment Strategy for Multi-vessel Coronary Artery Disease

**DOI:** 10.1101/2023.05.17.23290067

**Authors:** Harsh Rawal, Tung D Nguyen, Efehi Igbinomwanhia, Lloyd W Klein

## Abstract

**Introduction:** Fractional flow reserve (FFR) and instantaneous wave-free ratio (iFR) have become the standards for assessing physiological significance of intermediate-to-severe lesions on coronary angiography. However, the utilization of invasive physiology in patients with multi-vessel coronary artery disease (CAD) has not been definitively explored. The aim of this study is to examine how physiologic testing is applied to select treatment strategy in the clinic, and whether physiologic testing leads to differences in selection of treatment modality among patients presenting with two- and three-vessel CAD. The analysis identified how treatment strategy was decided based on FFR/iFR values in vessels selected clinically. Additionally, the differences in selection of treatment modality based on whether the vessel tested was the clinical target stenosis vessel was assessed.

**Methods:** In this series, 270 consecutive patients with angiographically determined multi-vessel disease who underwent FFR and/or iFR testing were included, and patients were classified initially based on their FFR/iFR results (normal or abnormal). Lesions were subsequently classified into target or non-target lesions based on clinical and non-invasive testing to identify if they were culprit lesions or not. Ordinal variables were analyzed using GraphPad Prism 8.4 (GraphPad Software). One-way ANOVA was used to determine if differences existed among the 3 subgroups prior to chi square testing. Chi-square goodness of fit test was used to determine if the treatment modality was chosen based on FFR/iFR values and lesion classification.

**Results:** FFR/iFR was abnormal in 51.9% of cases. Of the abnormal cases, 51.4% received coronary stenting (PCI) and 44.3% had bypass surgery (CABG). When the tested vessel was normal, CABG was least utilized. When the non-target vessel was tested, there was a substantial preference for PCI, however, when the target lesion was tested, all three treatments were used equally. In two-vessel CAD, the strategy depended on physiologic testing result of the target vessel. Medical therapy was preferred when the target vessel was normal, however, PCI was preferred if the target vessel was abnormal. In non-target vessel testing, PCI was preferred regardless of the physiologic testing result. The preferred treatment modality was substantially different in three-vessel CAD patients. Abnormal testing in target lesions was associated with significantly more CABG than medical therapy. PCI was preferred if the non-target vessel was tested. Furthermore, the incidence of tested lesions was significantly higher when located in the left anterior descending (LAD) when compared to other coronary arteries (*e.g.* left circumflex (LCX), right coronary artery (RCA)). In patients with two-vessel CAD, regardless of whether a target or non-target lesion was being tested, patients with LAD stenoses were more frequently treated by PCI.

**Conclusion:** The use of invasive physiologic testing in patients with multi-vessel CAD may alter the preferred treatment strategy. In clinical practice, this leads to a substantial increase in the use of PCI.

## Introduction

Fractional flow reserve (FFR) and instantaneous wave-free ratio (iFR) are the standards for assessing the physiological significance of anatomically ambiguous or intermediate lesions as well as angiographically significant lesions on coronary angiography. Deferral of revascularization based on normal FFR/iFR is cost effective and associated with good long-term clinical outcomes [1-3]. Functional assessment rather than anatomic estimates of severity is associated with improved long-term outcomes in intermediate lesions, improving clinical decision-making [4].

These adjunctive diagnostic procedures appropriately influence the decision for coronary revascularization, guide the performance of percutaneous coronary interventions (PCI), and optimize procedural outcomes. However, they are underutilized in contemporary practice, where the rate of FFR usage during PCI for intermediate coronary stenosis (40-70% diameter stenosis) is merely 6.1% [5]. In part, this low utilization is due to the additional time and cost of this additional invasive procedure, concerns about physician reimbursement, the potential for adverse events, and reluctance to employ the procedure unless it is going to yield additional treatment insights.

The optimal strategic application of invasive physiology in patients with multi-vessel coronary disease has not been definitively explored. Physiologic lesion evaluation is a powerful adjunctive diagnostic modality in the evaluation of intermediate severity coronary stenoses, but the factors governing its employment and its actual impact on clinical decision-making in this patient population is unknown and likely complex. Accordingly, we sought to characterize how these tests are used in practice.

## Methods

### Study objectives

This retrospective study was undertaken to determine how clinicians use invasive physiologic testing to select treatment strategy during catheterization. The distribution of patients whose treatment strategy was decided based on FFR/iFR values in vessel locations selected clinically was characterized. Additionally, differences in selection of treatment modality among patients presenting with two- and three-vessel CAD from their FFR/iFR results and whether the vessel tested was the clinical target stenosis vessel were assessed. Vessels were classified into target and non-target vessels based on pre-defined clinical criteria.

The primary objective of the study was to determine how FFR/iFR values are used to determine revascularization strategy in patients with angiographic multi-vessel CAD. The secondary objective of the study was to assess if selection of patients for physiologic testing showed any identifiable predispositions. The Institutional Review Board (IRB) approved the study, and all data were collected in a database that is encrypted and compliant with HIPAA regulations. All patient identifiers were kept confidential in compliant with HIPAA regulations.

### Eligibility

Over a 4-year period, 1541 consecutive patients underwent FFR/iFR at a single urban teaching hospital. Of these, 270 patients with angiographically determined multi-vessel (two- or three-vessel) disease underwent FFR and/or iFR testing and were eligible for entry to the study. Patients with previous CABG and significant left main stenoses were excluded from this study. The interventional cardiologist made the decision as to which vessel to test or not to test.

### Data acquisition and analysis

All relevant medical records and reports, including coronary angiograms, were reviewed. Retrospective registry data regarding admission diagnoses, emergency room documentation, past medical history, laboratory results (including troponin levels), cardiac stress test reports, cardiac angiogram reports, electrocardiograms, operative notes, and disposition at discharge were also compiled. Over 50 angiographic, clinical and physiologic variables were collected and evaluated to determine how these tests altered the initial treatment plan. Patient demographics (including age, gender, BMI, admitting diagnosis, indication for cath/PCI, and FFR/iFR) and other pertinent variables were acquired from data fields within the local NCDR Cath-PCI registry, including angiographically-determined stenosis severity and number of vessels diseased (based on angiographic description of ≥70% diameter stenosis).

### FFR and iFR parameters

FFR is the ratio of mean distal coronary pressure (P*_d_*) to mean aortic pressure (P*_a_*) during maximum hyperemia, which is usually induced by adenosine bolus or infusion, and represents the percentage of normal flow across a coronary stenosis. iFR is measured during a select portion of diastole, the wave-free period (WFP), when the forces that influence coronary flow are quiescent. iFR is calculated by measuring the resting trans-lesional pressure ratio (Pd/Pa) during the WFP. FFR and iFR values were considered abnormal if FFR≤0.80 or iFR≤0.89. Markedly abnormal values were considered if FFR<0.75 or iFR<0.85.

### Target lesion classification

Lesions were classified into target and non-target lesions by the clinicians who managed the cases. Target lesions were identified as the clinically apparent culprit lesion by morphology or severity, and believed clinically to be causing an acute coronary syndrome based on EKG or abnormal stress test. Non-target lesions were stenoses in other vessels identified by the angiogram to be intermediate or high severity but not directly related to the clinically affected area of the myocardium (*i.e.* in a contralateral vessel). Clinical correlation with ECG changes, results of the non-invasive evaluation, and other factors were used to make this determination, which was blinded to physiologic outcomes and revascularization strategy employed. Angiographic reports and clinical notes were reviewed to find the rationale behind selecting treatment strategy (*e.g.* medical treatment vs. PCI vs. CABG) as well as assessing lesion location by vessel (*e.g.* LAD vs. LCX vs. RCA vs. other vessels).

Patients with normal and abnormal FFR/iFR values were further subcategorized based on whether the target lesion was involved in the treatment and if treatment included other vessels. Each group was then subdivided based on the number of diseased vessels (*i.e.* two- vs. three-vessel CAD). Initial appraisal was performed regarding whether the lesions studied physiologically were the primary targets identified clinically or a secondary vessel, how many stenoses and diseased vessels were present and treated, and if the FFR/iFR outcomes were normal or abnormal. Age, gender, BMI, COPD, diabetes and other demographic variables were collected and were not independently predictive of the use of FFR/iFR in patients. No patient had revascularization (PCI or CABG) done in a vessel shown to have a normal FFR or iFR. When a revascularization was performed in a patient classified as having a normal test result, it was always performed in a different vessel than the one tested.

### Statistical analysis

Ordinal variables were analyzed using GraphPad Prism 8.4 (GraphPad Software) and presented as absolute values and percentages. One-way ANOVA was used to determine if differences existed among the 3 subgroups prior to chi square testing. Chi-square goodness of fit test was used to determine whether the treatment modality was chosen based on FFR/iFR values and lesion classification. The statistical analysis (chi-square) was run on every possible facet and combination with regards to FFR/iFR evaluations and number of diseased vessels. Moreover, chi-square goodness of fit is an ideal statistical choice for this study as the expected outcome frequency for each treatment modality in each patient group (two-vessel and three-vessel) is based on the FFR/iFR outcomes. Thus, chi-square analyses comparing the observed outcomes with the expected outcomes for each treatment modality in each patient group would allow the detection of biases (deviations from the expected) in the treatment modality chosen in different patient groups. A value of p<0.05 is considered significant.

## Results

A total of 270 patients with multi-vessel CAD who underwent FFR/iFR during coronary angiography or stenting comprised the entire study group. **Table 1** summarizes the treatment preferences based on angiographic extent of disease and the results of physiologic testing. Baseline patient characteristics (*e.g.* demographics, clinical characteristics, admitting diagnosis) are summarized in **Table 2**. Of the total number of patients, 113 (41.85%) underwent angiography due to NSTEMI/STEMI being their main indication, while 91 (33.70%) underwent the procedure due to an abnormal stress test (**Table 2**). The remaining patients had testing for malignant arrhythmias or for other causes, such as new onset cardiomyopathies, evaluation of worsening CHF, and valvular heart disease. There were no notable differences in baseline characteristics between patients with normal or abnormal FFR/iFR testing (**Table 2**). Similarly, there were no differences in baseline characteristics between patients receiving PCI, CABG, or medical treatment.

**Table 1.** Summary of results. Influence of angiographic extent of disease and result of physiologic testing on the preference of treatment modality.

| # of vessels diseased | Target or Non-target vessel tested | Normal FFR/iFR value | Abnormal FFR/iFR value |
| --- | --- | --- | --- |
| Two-vessel CAD | Target | Medical Mx | PCI |
|  | Non-target | PCI | PCI |
| Three-vessel CAD | Target | No trend | CABG |
|  | Non-target | PCI | PCI or CABG |

**Table 2.** Patient characteristics. Baseline characteristics of multi-vessel CAD patients who underwent FFR and/or IFR during coronary angiography and/or stenting. Patient baseline characteristics were further categorized based on FFR/iFR values (normal and abnormal) or selected treatment strategy (percutaneous coronary intervention (PCI), coronary artery bypass graft (CABG), and medical treatment (Med Mx)).

| Patient characteristics | Total (n=270) |  | Normal FFR/iFR (n=130) |  | Abnormal FFR/iFR (n=140) |  | p-value | PCI (n=132) |  | CABG (n=72) |  | Med Mx (n=66) |  | p-value |
| --- | --- | --- | --- | --- | --- | --- | --- | --- | --- | --- | --- | --- | --- | --- |
|  | # | % | # | % | # | % |  | # | % | # | % | # | % |  |
| Demographics |  |  |  |  |  |  |  |  |  |  |  |  |  |  |
| Age (SD) | 65 (10) | N/A | 65 (11) | N/A | 66 (10) | N/A | 0.861 | 65 (10) | N/A | 65 (9) | N/A | 66 (11) | N/A | 0.885 |
| Sex: Male | 207 | 76.7 | 101 | 48.8 | 106 | 51.2 | 0.781 | 102 | 49.3 | 58 | 28.0 | 47 | 22.7 | 0.554 |
| Race: White | 122 | 45.2 | 52 | 42.6 | 70 | 57.4 | 0.104 | 57 | 46.8 | 38 | 31.1 | 27 | 22.1 | 0.343 |
| Weight (SD) | 87 (19) | N/A | 87 (20) | N/A | 85 (19) | N/A | 0.808 | 89 (19) | N/A | 87 (18) | N/A | 85 (20) | N/A | 0.855 |
| BMI (SD) | 30 (7) | N/A | 31 (7) | N/A | 29 (7) | N/A | 0.796 | 31 (7) | N/A | 29 (6) | N/A | 29 (7) | N/A | 0.803 |
| Clinical characteristics |  |  |  |  |  |  |  |  |  |  |  |  |  |  |
| Diabetes mellitus | 129 | 47.8 | 60 | 46.5 | 69 | 53.5 | 0.428 | 71 | 55.0 | 32 | 24.8 | 26 | 20.2 | 0.498 |
| HbA1c (SD) of diabetics | N/A | 7.1 (1.9) | N/A | 7.3 (2.1) | N/A | 6.9 (1.7) | 0.769 | N/A | 7.0 (1.8) | N/A | 7.0 (1.5) | N/A | 7.3 (2.4) | 0.884 |
| Congestive heart failure | 58 | 21.5 | 28 | 48.3 | 30 | 51.7 | 0.793 | 27 | 46.6 | 17 | 29.3 | 14 | 24.1 | 0.724 |
| Previous PCI | 101 | 37.4 | 47 | 46.5 | 54 | 53.5 | 0.486 | 45 | 44.6 | 30 | 29.7 | 26 | 25.7 | 0.561 |
| Admitting diagnosis |  |  |  |  |  |  |  |  |  |  |  |  |  |  |
| NSTEMI/STEMI | 113 | 41.9 | 58 | 51.3 | 55 | 48.7 | 0.778 | 58 | 51.3 | 25 | 22.2 | 30 | 26.5 | 0.804 |
| Positive stress test | 91 | 33.7 | 43 | 47.3 | 48 | 52.7 | 0.753 | 43 | 47.3 | 28 | 30.8 | 20 | 21.9 | 0.478 |
| Arrhythmia | 6 | 2.2 | 2 | 33.3 | 4 | 66.7 | 0.414 | 3 | 50.0 | 2 | 33.3 | 1 | 16.7 | 0.846 |
| Others | 60 | 22.2 | 27 | 45.0 | 33 | 55.0 | 0.439 | 28 | 46.7 | 17 | 28.3 | 15 | 25.0 | 0.718 |

140 patients (51.9%) had abnormal FFR/iFR testing of at least one lesion, while the remaining 130 patients (48.1%) had normal FFR/iFR testing (**Fig. 1**). Of the patients with abnormal FFR/iFR testing, very few were treated medically (6 patients, 4.3%) when compared to either PCI (72 patients, 51.4%) or CABG (62 patients, 44.3%) (**Fig. 2A**). Of the patients with normal FFR/iFR, substantially more were treated medically or with PCI (60 patients, 46.2%) than with CABG (10 patients, 7.5%) (**Fig. 2A**). Thus, abnormal physiologic assessment led to overall increased revascularization and decreased medical management.

**Figure 1.**
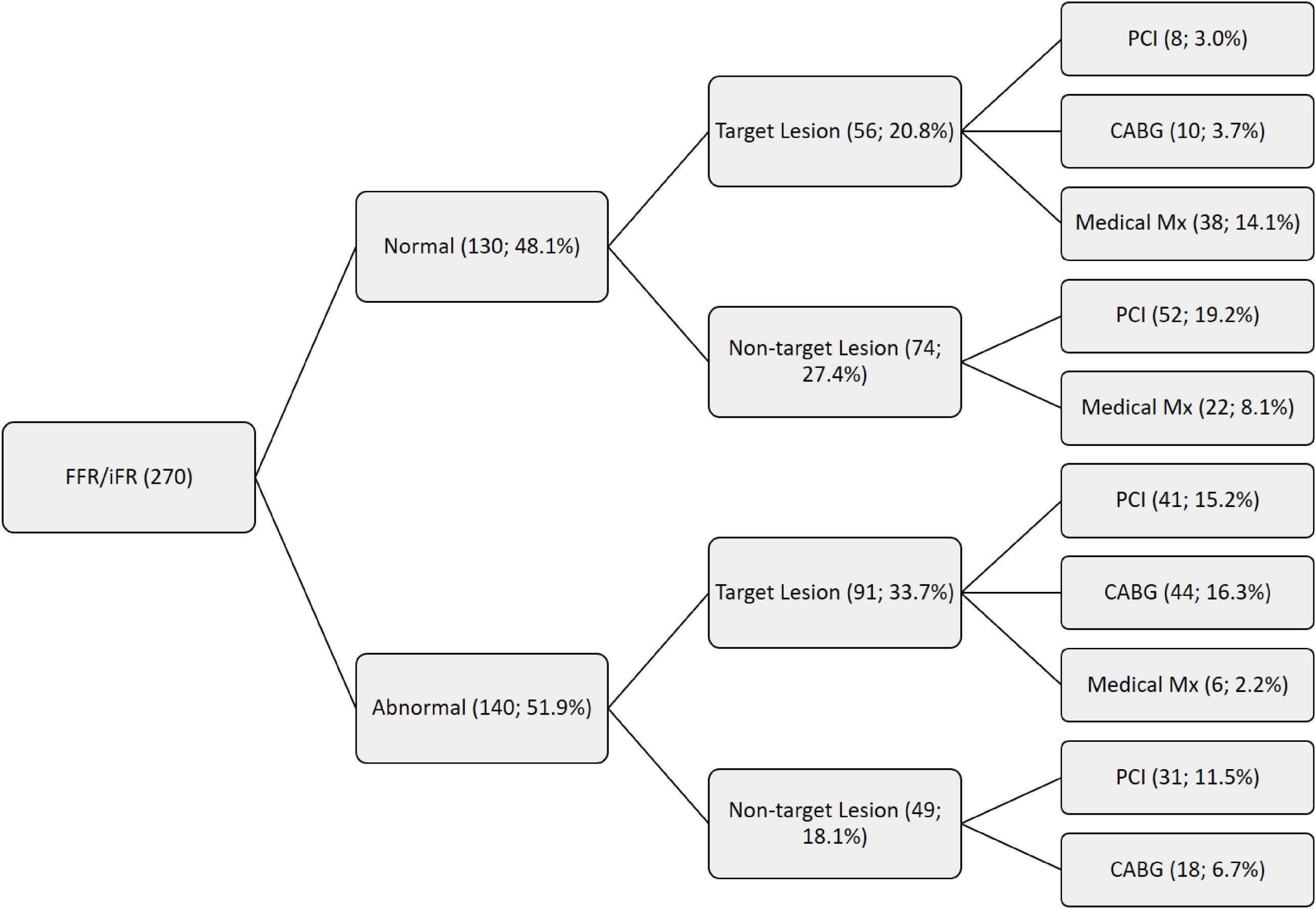
Patient cohort with angiographically-determined multi-vessel coronary artery disease and underwent FFR and/or iFR evaluations.

**Figure 2.**
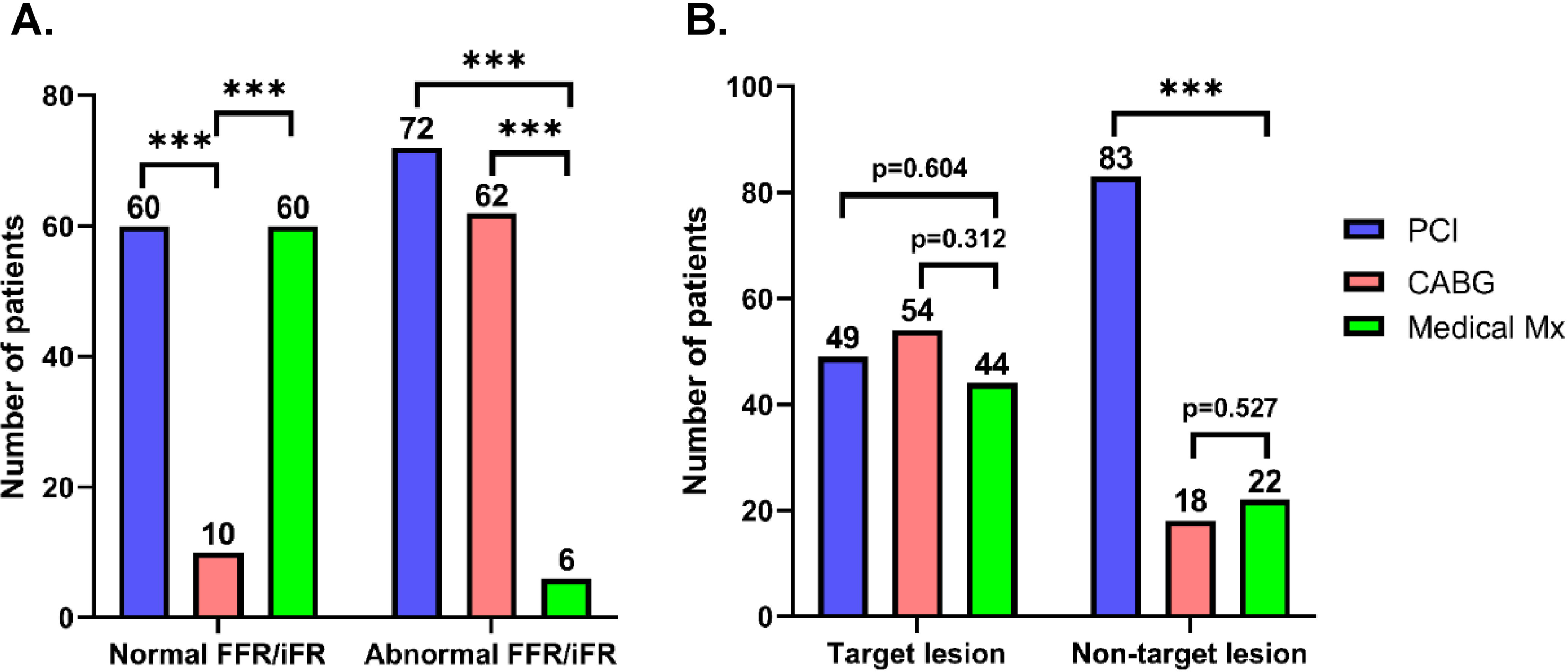
Overall proportions of treatment modalities and lesion locations by FFR/iFR results. Patients were categorized based on the selected treatment strategy (percutaneous coronary intervention (PCI), coronary artery bypass graft (CABG), and medical treatment (Med Mx)) as well as by FFR/iFR values (normal and abnormal) **(A)** or lesion types (target and non-target) **(B).** *** indicates p<0.001.

The relationship between the lesions selected for invasive physiologic measures (target vs. non-target) and the subsequent treatment strategy was also assessed. Of the 123 patients with non-target lesions tested, significantly more patients received PCI (83 patients, 67.5%) than were treated medically (22 patients, 17.9%) (**Fig. 2B**). However, there was not a notable difference between the number of patients who underwent bypass surgery (18 patients, 14.6%) and the number of patients treated medically (**Fig. 2B**). This skew was not observed with the treatment employed in the 147 patients with target lesions tested. There were no differences in the distribution of patients receiving medical therapy (44 patients, 30.0%) compared to patients receiving PCI (49 patients, 33.3%) or undergoing CABG (54 patients, 36.7%) (**Fig. 2B**).

Patients were also examined based on the lesion location by vessel. In normal FFR/iFR group, the majority of the lesions were located in the LAD (49 patients, 37.7%), followed by LCX (31 patients, 23.8%), RCA (28 patients, 21.5%), and lastly the other branches (22 patients, 16.9%) (**Fig. 3A**). The distribution of lesions in patients with abnormal FFR/iFR highlighted lesions predominant in the LAD (96 patients, 68.6%) (**Fig. 3A**). Similarly, analyses of both target and non-target lesions showed substantially more lesions located in the LAD (52.4% and 54.5%, respectively) compared to the LCX, RCA, or other branches (**Fig. 3B**). Overall, lesions were primarily located in the LAD regardless of physiologic assessment values or lesion classification.

**Figure 3.**
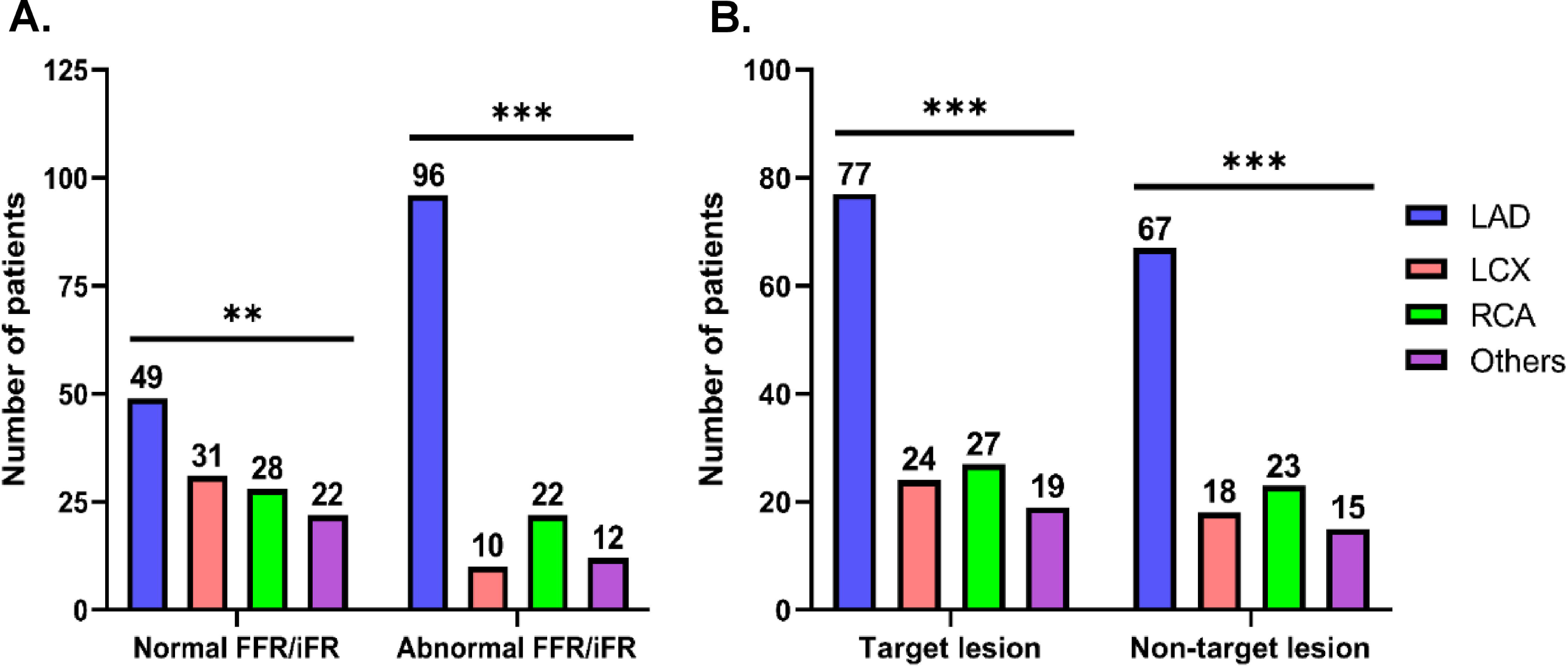
Overall proportions of treatment modalities and lesion locations by lesion classification. Patients were categorized based on the location of the lesion involved in the treatment (left anterior descending (LAD), left circumflex (LCX), right coronary artery (RCA), and other vessels) as well as by FFR/iFR values (normal and abnormal) **(A)** or lesion types (target and non-target) **(B).** ** indicates p<0.01, and *** indicates p<0.001.

### Abnormal physiologic assessment leads to increased target and non-target lesion revascularization in two-vessel CAD

149 patients with two-vessel CAD had undergone FFR/iFR testing. 44 patients with two-vessel CAD had normal FFR/iFR testing in the target lesion, and the majority were treated medically (32 patients, 72.6%) compared to either PCI or CABG (6 patients, 13.8%) (**Fig. 4A**). Intriguingly, substantially more patients with abnormal FFR/iFR in the target lesion received PCI (29 patients, 78.4%) and CABG (7 patients, 18.9%) than medical therapy (1 patient, 2.7%) (**Fig. 4A**). These results highlight significant preference for revascularization in patients presenting with two-vessel CAD and abnormal FFR/iFR testing in target lesions. 44 patients with two-vessel CAD patients had normal FFR/iFR testing in non-target lesions, and interestingly the majority received PCI (35 patients, 79.5%) over medical therapy (9 patients, 20.5%), with no patients undergoing CABG (**Fig. 4B**). Conversely, significantly more patients with abnormal FFR/iFR in non-target lesions received PCI (18 patients, 75.0%) than CABG (6 patients, 25.0%), and no patients were treated medically (**Fig. 4B**).

**Figure 4.**
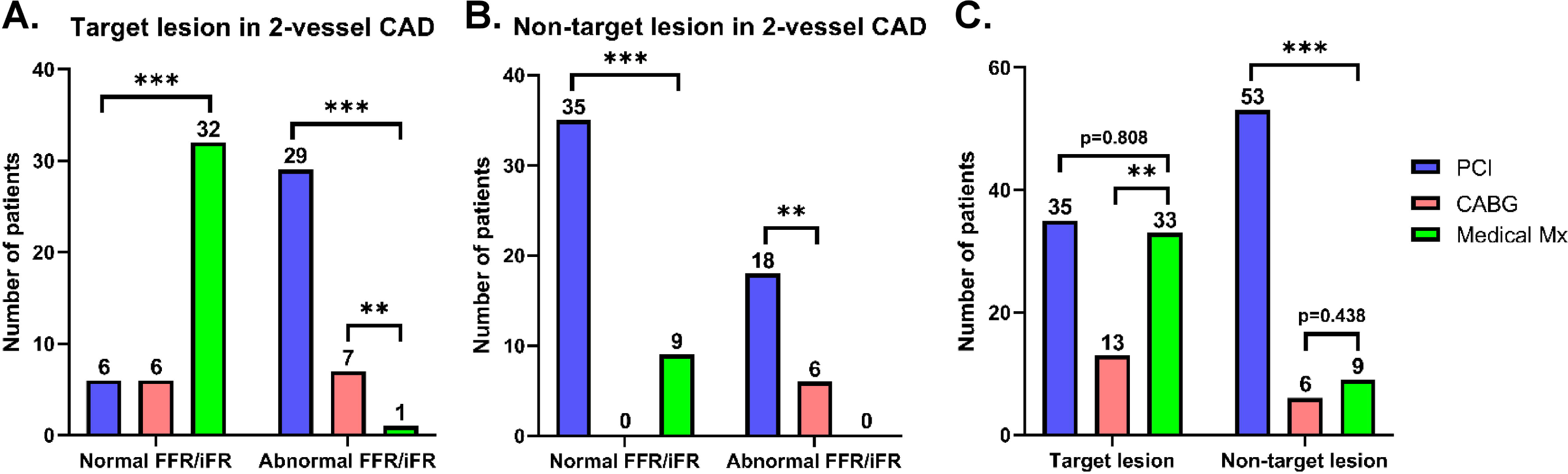
Proportions of treatment modalities and lesion locations by FFR/iFR results in two-vessel CAD. Two-vessel CAD patients were categorized by the selected treatment strategy (percutaneous coronary intervention (PCI), coronary artery bypass graft (CABG), and medical treatment (Med Mx)) in addition to FFR/iFR values (normal and abnormal). Patients were further subcategorized based on whether the target lesion was involved in the treatment **(A)** or if the treatment included other vessels **(B).** These patients were then separated based on the selected treatment strategy and lesion types (target and non-target) regardless of FFR/iFR results **(C).** ** indicates p<0.01, and *** indicates p<0.001.

68 two-vessel CAD patients with non-target lesions were tested, and the majority received PCI (53 patients, 78.0%) instead of medical therapy (9 patients, 13.2%) or CABG (6 patients, 8.8%) (**Fig. 4C**). 81 two-vessel CAD patients with target lesions were tested, and significantly more patients were treated medically (33 patients, 40.7%) or with PCI (35 patients, 43.2%) than with CABG (13 patients, 16.1%) (**Fig. 4C**). Thus, with two-vessel CAD patients, abnormal FFR/iFR testing in either target or non-target stenoses led to an overall increase in revascularization (PCI or CABG) and decrease in medical management. The skew favoring PCI was especially prevalent when testing non-target lesions in two-vessel CAD patients, where substantially more PCIs were performed than medical management or CABG regardless of FFR/iFR results.

When examining the lesion distribution in target vessels of two-vessel CAD patients, we found significantly higher lesion counts in the LAD than in other arteries, which were seen with normal and abnormal FFR/iFR testing (**Fig. 5A**). Similar observations were found regarding lesion distribution in non-target vessels of two-vessel CAD patients (**Fig. 5B**). Lastly, in patients with two-vessel CAD regardless of whether a target or non-target lesion was being tested, lesion counts were significantly higher in LAD than in other arteries (**Fig. 5C**). Taken together, this strongly suggests that two-vessel CAD patients with LAD stenosis were more frequently treated by PCI.

**Figure 5.**
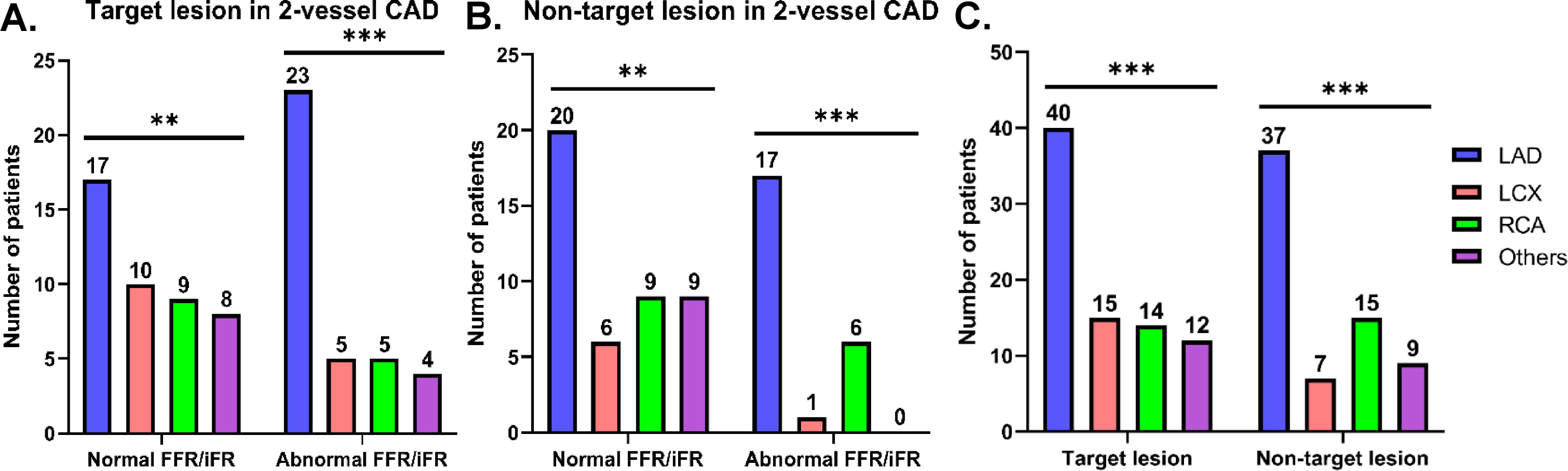
Proportions of treatment modalities and lesion locations by lesion classification in two-vessel CAD. Two-vessel CAD patients were categorized by the location of the lesion involved in the treatment (left anterior descending (LAD), left circumflex (LCX), right coronary artery (RCA), and other vessels) in addition to FFR/iFR values. Patients were further subcategorized based on whether the target lesion was involved in the treatment **(A)** or if the treatment included other vessels **(B).** These patients were then separated based on the lesion location and lesion types (target and non-target) regardless of FFR/iFR results **(C).** ** indicates p<0.01, and *** indicates p<0.001.

### Abnormal physiologic assessment leads to increased target and non-target lesion revascularization in three-vessel CAD

121 patients with three-vessel CAD had undergone FFR/iFR testing. 54 patients with three-vessel CAD had abnormal FFR/iFR values in the target lesion, and the majority underwent CABG (37 patients, 68.5%) instead of PCI (12 patients, 22.2%) or medical treatment (5 patients, 9.3%) (**Fig. 6A**). Conversely, of the patients with normal FFR/iFR testing in the target lesion, there were no notable differences between the number of patients treated medically (6 patients, 50.0%) compared to those who received PCI (2 patients, 16.7%) or CABG (4 patients, 33.3%) (**Fig. 6A**). These results highlight a preference towards CABG with abnormal FFR/iFR values in the target vessel in patients with three-vessel CAD. 25 patients with three-vessel CAD had abnormal testing in non-target lesions, and patients either received PCI (13 patients, 52.0%) or underwent CABG (12 patients, 48.0%) (**Fig. 6B**). Intriguingly, patients with normal FFR/iFR in non-target lesions either received PCI (17 patients, 56.7%) or were treated medically (13 patients, 43.3%), and no patients underwent CABG (**Fig. 6B**).

**Figure 6.**
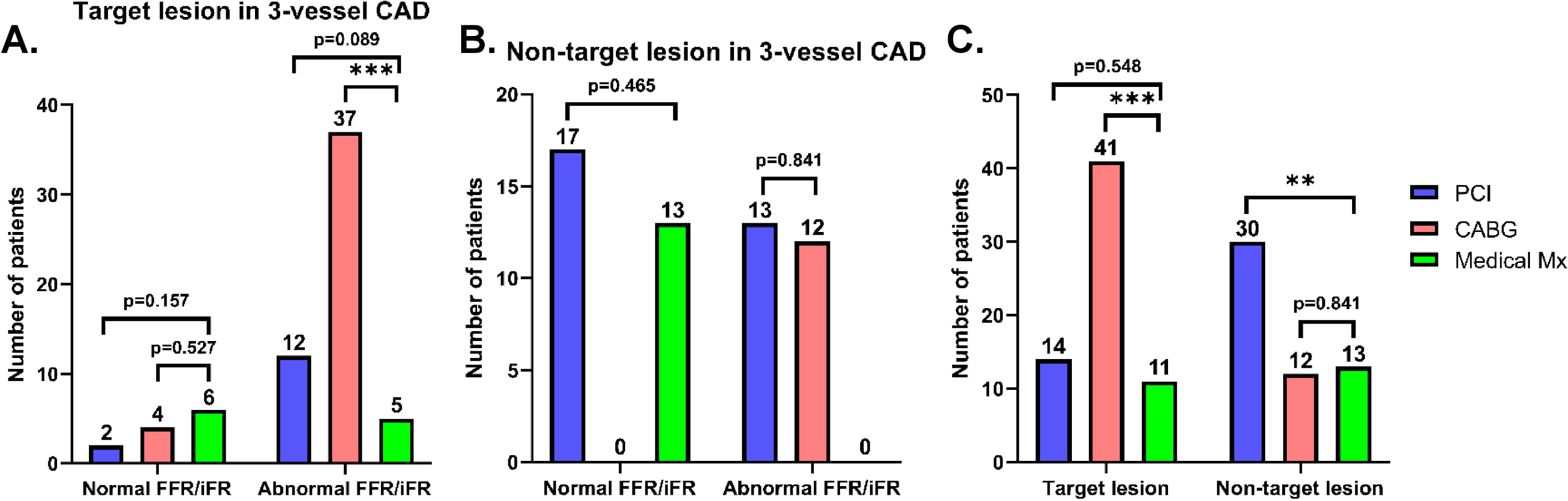
Proportions of treatment modalities and lesion locations by FFR/iFR results in three-vessel CAD. Three-vessel CAD patients were categorized by the selected treatment strategy (percutaneous coronary intervention (PCI), coronary artery bypass graft (CABG), and medical treatment (Med Mx)) in addition to FFR/iFR values (normal and abnormal). Patients were further subcategorized based on whether the target lesion was involved in the treatment **(A)** or if the treatment included other vessels **(B).** These patients were then separated based on the selected treatment strategy and lesion types (target and non-target) regardless of FFR/iFR results **(C).** ** indicates p<0.01, and *** indicates p<0.001.

55 three-vessel CAD patients with non-target lesions were tested, and the majority received PCI (30 patients, 54.6%) instead of medical therapy (13 patients, 23.6%) or CABG (12 patients, 21.8%) (**Fig. 6C**). 66 three-vessel CAD patients with target lesions were tested, and significantly more patients underwent CABG (41 patients, 62.1%) than medical treatment (11 patients, 16.7%) or with PCI (14 patients, 21.2%) (**Fig. 6C**). Our finding strongly suggests that FFR/iFR testing was employed in non-target vessels to determine which patients could be treated with PCI. If PCI could not be performed, then either CABG or medical therapy was chosen, likely depending on their candidacy based on non-angiographic criteria. Moreover, the bias against PCI was especially profound in three-vessel CAD patients whose target vessel was abnormal (42 vs. 12).

When examining the lesion distribution in three-vessel CAD patients with abnormal FFR/iFR testing, we found significantly higher lesion counts in the LAD than in other arteries, which were seen with both target and non-target lesions (**Fig. 7A-B**). Conversely, no notable differences were observed regarding lesion distribution in three-vessel CAD patients with normal FFR/iFR testing in either target or non-target lesions (**Fig. 7A-B**). Lastly, in patients with three-vessel CAD regardless of whether a target or non-target lesion was being tested, lesions counts were significantly higher in LAD than in other arteries (**Fig. 7C**). Overall, this strongly suggests that three-vessel CAD patients with abnormal testing and LAD stenosis were more frequently treated by either PCI or CABG, while no definitive inferences could be made regarding patients with normal testing.

**Figure 7.**
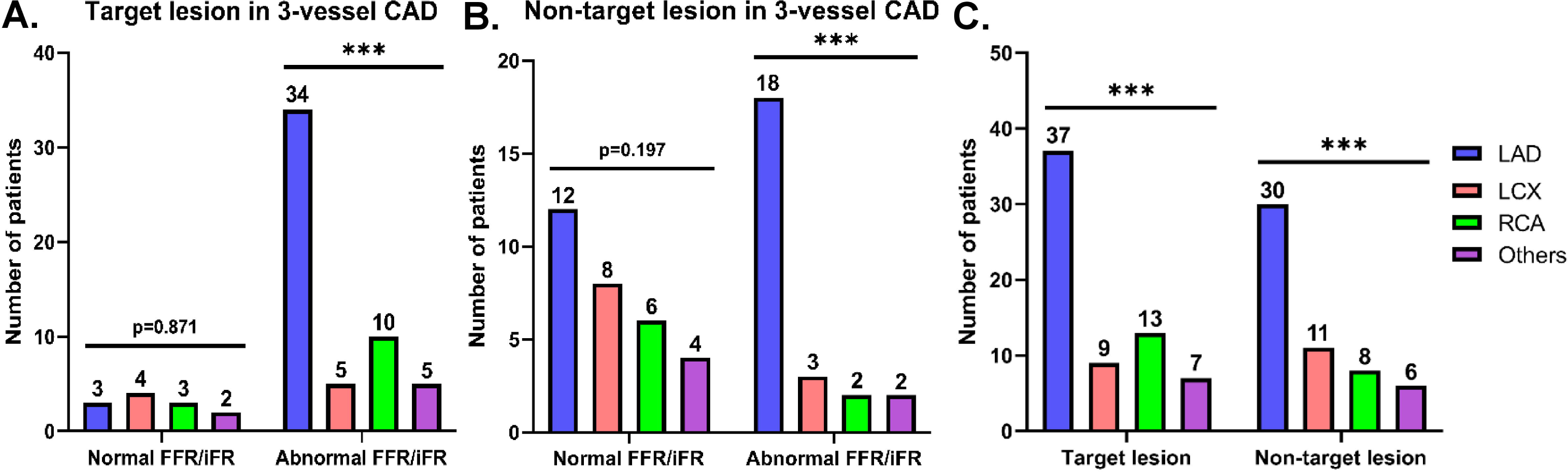
Proportions of treatment modalities and lesion locations by lesion classification in three-vessel CAD. Three-vessel CAD patients were categorized by the location of the lesion involved in the treatment (left anterior descending (LAD), left circumflex (LCX), right coronary artery (RCA), and other vessels) in addition to FFR/iFR values. Patients were further subcategorized based on whether the target lesion was involved in the treatment **(A)** or if the treatment included other vessels **(B).** These patients were then separated based on the lesion location and lesion types (target and non-target) regardless of FFR/iFR results **(C).** *** indicates p<0.001.

## Discussion

FFR/iFR testing is the standard invasive assessment of physiologic impact of lesions on the myocardium in coronary angiography. The aims of this study were to examine how physiologic testing is applied in selecting treatment strategy and to evaluate differences in selection of treatment modality among patients presenting with two- and three-vessel CAD. The analysis identified how treatment strategy was decided based on FFR/iFR outcomes in vessel locations selected clinically. Differences in the selection of treatment modality based on whether the vessel tested was the clinical target stenosis vessel were also assessed.

These results demonstrate the use of functional revascularization in contemporary practice. Affected vessels are chosen for testing when proving the severity of a lesion in a particular vessel will alter the optimal strategy. In real world contemporary practice, invasive physiological testing is not applied randomly, but rather it is used in focused situations when ascertaining the functional number of vessels diseased may alter the preferred treatment strategy. In most situations, this resulted in increased PCI as the treatment strategy. This skew can be identified by case selection for these tests, as the treatment modality changes based on the lesion classification (target vs. non-target) as well as the number of diseased vessels involved (two-vessel vs. three-vessel). While the nature of the bias is predicated on finding the best treatment strategy according to the evidence base and by practice guidelines, the use of physiologic testing might lead to increased PCI use in multi-vessel CAD.

Since the introduction of FFR in 1993 [6] and most recently iFR in 2012[7], physiology-guided revascularization has provided evidence-based approach in managing patients with CAD. Multiple landmark trials [8, 10] have shown that FFR-guided revascularization strategy to be safe, cost effective and associated with reduced adverse cardiac events compared to angiography-guided strategy. Non-inferiority of iFR to FFR in predicting cardiovascular outcomes and a reasonable alternative for physiologic assessment has also been demonstrated. In patients with multi-vessel disease, non-invasive testing often underestimates the burden of coronary stenosis [11]. Even though non-invasive tests, such as stress echocardiography and myocardial perfusion scintigraphy, provide information regarding ischemic burden, these tests have poor discrimination in identifying which lesions cause ischemia, especially in patients with multi-vessel disease (11). Hence, there is a strong need for accurate functional testing to identify if a specific coronary lesion produces ischemia or not. FFR guided complete revascularization was associated with a significant 12.7% absolute reduction in major adverse cardiovascular events, mainly due to fewer revascularizations in the FFR group [12]. This could lead to a paradigm shift in the indications for revascularization in patients with stable CAD. Physiologic measures prior to deciding on revascularization may decrease the rate of implanted stents or inserted grafts. It could also change the patient classification according to number of diseased vessels, perhaps downgrading patient classification if the stenoses are not functionally significant and thus increasing the use of PCI.

About 70% of patients referred for PCI have multi-vessel CAD [13], and in these patients non-invasive testing is often unreliable. FFR/iFR can be used to aid decision-making, especially in cases where there is discordance between lesion severity/location and non-invasive testing. The FAME-2 trial demonstrated the superiority of FFR-guided PCI plus medical therapy over medical management alone in patients with multi-vessel CAD. The FFR-guided group showed lower rates of the primary endpoint of all-cause death, non-fatal MI, and repeat revascularization. This difference was largely driven by a greater need for urgent revascularization in patients managed with medical therapy alone [14]. These findings raise curious questions about the utility of physiologic parameters in selection of patients for CABG, where the anticipation that non-significance in one or more vessels would prevent a patient from an open-heart surgery and instead receive a coronary stent. A study led by Fearon et al [15] observed about 1500 patients from 48 centers, and they found that in patients with CAD, FFR-guided PCI was not found to be inferior to CABG with regards to incidence of death, MI, stroke or repeat vascularization at 1-year.

Multiple trials including SYNTAX, SYNTAX II l, FAME, and FAME II have evaluated the impact and use of FFR on PCI and revascularization. FAME demonstrated lower rates of major adverse cardiac and cerebrovascular events with use of FFR. FAME II also used FFR guided modality for revascularization in combination with optimal medical therapy, and their outcomes demonstrated a benefit against patient mortality with physiologic revascularization [2]. A recent meta-analysis analyzed the prognostic value of FFR, where linking physiologic severity to clinical outcomes suggested that FFR-guided strategy leads to revascularization approximately half as often as anatomic-based strategies [16]. FFR-guided strategy was also associated with 20% lesser incidence of adverse events and 10% better angina relief [16]. An evolving discussion regarding the appropriateness of stenting a lesion is based on its functionality that may lead to significant occlusion. FFR can help us distinguish lesions, which are functionally significant and obstructive, from non-obstructive lesions, and thus assist in guiding treatment selection when choosing PCI vs. bypass graft [17].

Society guidelines summarize the value of physiologic testing in appropriate treatment selection for patients to direct them for either PCI or CABG [18]. The functional syntax score will differ based on angiographic interpretation and inclusion of intermediate but non-ischemic lesions. This might lead to reclassification of an angiographic three-vessel CAD to one- or two-vessel CAD, which would therefore benefit from PCI rather than CABG. A direct comparison between the outcomes of SYNTAX II and SYNTAX I were performed, where iFR was performed in 74% of lesions and consequently led to deferring treatment in 31% of interrogated lesions. At 1-year, PCI with this treatment strategy had similar outcomes to the CABG cohort in SYNTAX I. The PCI cohort, however, had lower MACE scores compared to SYNTAX-I PCI cohort (HR 0.58 (95% I 0.39-0.85). The two-year follow up for this study is still pending [18].

This study is ancillary to the FAME 3 trial, which investigated whether an FFR-guided PCI is non-inferior to CABG in patients with multi-vessel CAD. As such, these observations may help determine if the use of these physiologic tests in contemporary practice beyond the abovementioned studies is appropriate. Long-term data on the effects of CABG on angiographically-borderline stenoses are still unknown. There are data to support that there was no difference in MACE at 3-years if patients underwent CABG vs. FFR guided PCI [19]. These data further confirm that transition towards physiologic assessment with FFR/iFR, which gives more objective evidence of the severity of a lesion, should be given key importance in clinical decision-making. This is especially important in lesions where there are no stress tests available or the results of the anatomic lesions and stress tests are discordant [17].

Recently, the RIPCORD2 trial discussed blanket use of FFR as a diagnostic tool for all patients undergoing coronary angiography. In the 1100-patient study, there was no significant difference between in-hospital costs and quality of life at 1-year when testing with angiography alone when compared to angiography plus FFR usage. Interestingly, routine FFR did not reduce costs, improve quality of life or reduce major adverse cardiac events or revascularization rates compared to angiography alone. Conversely, it was associated with higher complication rates, contrast use, and procedural times [20].

There have been various studies assessing the use of FFR in the context of CABG selection. Spadaccio et al [21] accounted for all the different trials conducted worldwide on the subject. The conclusions were based on the mechanisms of flow competence and whether the grafts used were arterial or venous. This study concluded that preoperative use of FFR reduces the number of distal anastomoses and simplifies CABG procedure, however, the data on improved early clinical outcomes were limited. Furthermore, preoperative data showed some correlation with arterial grafts but evidence of FFR usage in venous grafts was lacking. While the use of FFR in surgical revascularization is not definitively explored, there are some benefits of physiological testing in deciding treatment strategy [21].

The main limitation of the current study is that it was a single center, non-randomized observational study. The data accurately reflects the use of these tests in current practice, however, several of the subgroups had too few patients to be certain that the lack of significant differences seen would be verifiable in a larger population. Angiographic reporting of stenosis severity is known to be subjective and inaccurate. Without a doubt, the angiographer had a sense of severity before FFR/iFR usage, and there is no way to retroactively determine how the physiologic tests impacted the percent stenosis reported.

In conclusion, despite the use of invasive physiologic testing in patients with multi-vessel coronary artery disease, the functional number of diseased vessels may alter the preferred treatment strategy. In most situations, this may lead to a substantial increase in the use of PCI as the treatment strategy.

## CONFLICT OF INTEREST DISCLOSURES

The authors declare that there is no conflict of interest regarding the publication of this paper. There was no funding associated with the study.

## ETHICS DISCLOSURES

This study has been approved by the human research ethics committee of the Advocate Health Care Network (Ref. No. 1243433-1).

## Data Availability

All data produced in the present study are available upon reasonable request to the authors.

## Notes

### Competing Interest Statement

The authors have declared no competing interest.

